# In search for the hotspots of Disease X: A biogeographic approach to mapping the predictive risk of WHO’s Blueprint Priority Diseases

**DOI:** 10.1101/2020.03.27.20044156

**Authors:** Soushieta Jagadesh, Marine Combe, Mathieu Nacher, Rodolphe Elie Gozlan

## Abstract

Anthropization of natural habitats including climate change along with overpopulation and global travel have been contributing to emerging infectious diseases outbreaks. The recent COVID-19 outbreak in Wuhan, highlights such threats to human health, social stability and global trade and economy. We used species distribution modelling and environmental data from satellite imagery to model Blueprint Priority Diseases occurrences. We constructed classical regression and Support Vector Machine models based on environmental predictor variables such as landscape, tree cover loss, climatic covariates. Models were evaluated and a weighed mean was used to map the predictive risk of disease emergence. We mapped the predictive risk for filovirus, Nipah, Rift Valley Fever and coronavirus diseases. Elevation, tree cover loss and climatic covariates were found to significant factors influencing disease emergence. We also showed the relevance of disease biogeography and in the identification potential hotspots for Disease X in regions in Uganda and China.

**Article Summary Line:** In our study with the use of a biogeographic approach, we were able to identify Wuhan as a potential hotspot of disease emergence in the absence of COVID-19 data and we confirm that distribution of disease emergence in humans is spatially dependent on environmental factors.

## Introduction

Changes to local land use and biodiversity, the increasing international connectivity through human transport and trade and climate change provide optimal conditions for the emergence of zoonotic infectious diseases. The displacement of the geographical footprint of pathogens and/or infected hosts leads to such EIDs (*1*) with the COVID-19 as an ongoing example at the center of international scrutiny. The World Health Organization (WHO) has developed identify list of zoonotic diseases posing of a large-scale public health risk due to 1) their epidemic potential and 2) current absence or limited number of treatment and control measures available (*2*). The current list, established in 2018, includes Crimean-Congo hemorrhagic fever (CCHF), Ebola virus and Marburg virus disease (EVD & MVD), Lassa fever (LF), Middle East respiratory syndrome coronavirus (MERS-CoV) and Severe Acute Respiratory Syndrome (SARS), Nipah and henipaviral diseases, Rift Valley fever (RVF), Zika and Disease X. The term “Disease X” refers to a potential international epidemic caused by a currently unknown pathogen, for which WHO calls for preparedness in the event of a novel disease emergence. This list of diseases in need of urgent research was termed as Blueprint list of Priority Diseases (BPDs).

All the BPDs in the current list can be classified as zoonotic viral infections. In the past four decades, over 70% of the emerging infections are or were zoonoses (*3,4*). The increasing unpredictability in the global climate and the decreasing distance between the local human-animal-ecosystem interactions play a major role in driving the emergence of infection in human populations. The rising mean temperatures have been reported to have a significant effect in the tick-borne CCHF incidence and mosquito-borne Zika sustainability in subtropical and temperate regions (*5,6*). Bushmeat consumption and animal trading, arising from the growing demand of animal protein, causes significant changes at the human-animal reservoir interface (*7*). Among the BPDs, studies demonstrate that the SARS and EVD outbreaks were directly linked to the consumption of infected bushmeat (*8,9*). LF, MVD and EVD flourish in West and central Africa where the consumption of bushmeat is four times greater than the Amazon, which is richer in biodiversity (*10*). Moreover, EIDs are also triggered by anthropogenic pressures on land use for agricultural expansion and livestock farming to meet the demand of a growing human population. The fruit-bat migration driven by the deforestation through forest fires in the islands of Sumatra lead to the emergence of Nipah disease in farmers and abattoir workers in Malaysia (*11,12*). Once sufficient infection cycles between human-animal without the sustainable transmission among humans termed as viral chatter is established, the emergence of human-to-human transmission is inevitable (*7*). Mathematical modeling and prediction provides quick assessment for control and potential preventive efforts when time for epidemiological studies is scarce (*13*). Modeling of infection dynamics analyses diseases outbreaks in animal population and estimates the rate of transmission as well as the potential chance of spillover. Yet, the increasing trends of EIDs risks surpass our capacity in the surveillance and detection of spillovers and outbreaks. Thus, the current public health response to EIDs is to “get ahead of the curve” and the growing resolution of satellite imagery has shifted the paradigm towards identifying potential environmental drivers such as deforestation, land fragmentation, biodiversity loss and climate change rather than the surveillance of the EIDs themselves.

Here we proposed mapping the predictive risk of the BPDs using Species Distribution Models (SDM) based on potential environmental drivers such as deforestation, landscape and climatic covariates derived from satellite imagery following the year 2000. The aim is to provide a global perspective, measure predictive risks and evaluate the use of biogeography on predicting diseases outbreaks. We also used our approach to EIDs using disease biogeography as a tool to identify the potential hotspots for an unknown Disease X listed in the BPDs.

## Methods

To construct robust models and generate predictive risk data of the BPDs’ distribution, we adapted the following steps from species distribution modelling: 1) compilation of the geographical coordinates of the BPDs emergence; 2) construction of spatial buffer around the spatial points; 3) generation of pseudo absence points; 4) extraction of environmental predictor covariates; 5) fitting of the model and 6) calculation of the predictive risk of each BPD.

### Location of BPD emergence

We extracted the distributional data on the global occurrence of the BPDs in humans across the years 2000 to 2019 from WHO archives, Promed mail and published studies (Appendix Table.1). In cases where the origin of the BPDs were unclear, we narrow down to the general region or district of origin. In other words, the localization of BPDs emergence correspond to patient zero of each outbreak. We did not include the diseases endemic to countries such as CCHF and Lassa fever. The calculation of predictive risk for endemic diseases and pandemics such as Zika may not contribute to the existing evidence. Laboratory outbreaks and outbreak in domestic and wildlife were also excluded. We georeferenced the sites of origin or the centroids of the region of occurrence to the nearest 0.0001°. Spatial buffers were constructed around the geographical coordinates depending upon the mobility range of the respective pathogens reservoirs and intermediate buffers (Appendix table 2). The construction of buffers also mitigates the relatively less precise geographical logical coordinates in cases where the exact point of origin was unclear.

### Presence and Pseudo-absence points

We constructed spatial buffers to mitigate any potential inaccuracies in the localization of the occurrence points. For the presence points, we generated 100 random points at a 100km spatial buffer around the point of origin of each BPDs outbreak. Then, a spatial bufferzone of 150km was used as a mask and we generated 1000 pseudo-absence points within the geographical extent of the reservoirs of each of the BPDs. We chose the buffer of radius 150km to account to the average flying range of the order Chiroptera, the reservoirs of most BPD.

### Environmental predictor datasets

We extracted the climatic covariates such as monthly maximum and minimum temperatures and precipitation from ‘Terra Climate’ high-resolution global data from 2000-2017(*14*). Land cover data used was a composite of the 2017 MODIS satellite imagery and the deforestations mdata was derived from the Hansan’s deforestation tree loss-year maps from 2000-2018. The topological data including altitude and hydrological models was extracted from one arc-second digital elevation model (DEM) of 30m resolution, derived from NASA Version 3.0 Shuttle Radar Topography Mission (SRTM) imagery available in the United States Geological Survey (USGS). We obtained the geographical distribution of the primary hosts and reservoir mammals from the IUCN red list(*15*). The shapefiles were rasterized and resampled into rasters of the 5km resolution raster model. We used livestock distribution rasters (2010) from the Food and Agriculture Organisation (FAO) for diseases such as RVF and Nipah including cattle, goats and pigs (*16*). The raster layers were resampled to a fixed resolution of 5km and stacked to a raster brick.

### Fitting of model and Model prediction

The disease distribution models were fitted using the r package “discmo”. In our study, we used the main two methods for species distribution modelling (SDM) i) the classical generalized linear models (glm) using gaussian regression methods and ii) the machine learning method, support vector machine (SVM). We choose the glm models to analyze the influence of the environmental factors on the emergence of BPDs. SVM are popular in SDM using presence and pseudo-absence data. The models were evaluated using ROC curves and the area under curves (AUC) for the produced thresholds were calculated. Studies have critized the use of AUCs in the evaluation of SDMs, especially when the study involves large extent. We removed the “spatial sorting bias” through “point-wise distance sampling” as explained by Hijmanns, 2012. Model prediction was made using the “predict” function to map the predictive risk of the diseases based on the values of the independent variables extracted from the environmental predictor rasters. A weighed average of the glm and SVM models were calculated using the AUCs for each BPD model and a final composite prediction was made.

### Deforestation analysis

To analyze the impact of deforestation on the emergence of the BPDs, we conducted a detailed spatiotemporal analysis using the R package “gfcanalytics” on the Hansan’s tree cover loss maps. We used the presence points at a spatial buffer of 100km, used for the BPD models, for this analysis.

## Results

We modelled five out of nine BPDs. We used glm and SVM methods of SDM for each of the disease. The AUC was maintained at an average of 0.9323 for glm models and 0.9597 for the SVM models. We have tabulated the AUC for each model in Table 1.

**Table 1:** Generalized linear models (GLM) and support vector machine (SVM) regression coefficients for the AUC of the blue print diseases list analysis (BPD).

| **Disease/Model** | **Gaussian/GLM regression** | **Support Vector Machine** |
| --- | --- | --- |
| Ebola virus (EVD) | 0.9115 | 0.8680 |
| Marburg virus disease (MVD) | 0.9885 | 0.9913 |
| Coronavirus (CoV) | 0.8625 | 0.9831 |
| Nipah and henipaviral diseases | 0.9811 | 0.9911 |
| Rift Valley fever (RVF) | 0.9190 | 0.9653 |

### Filovirus diseases

The distribution of the filovirus diseases, EVD and MVD, was restricted to the African subcontinent (Figures 1 & 2). The climatic covariates such as average total monthly precipitation [95%CI 0.0091 to 0.0305; P 0.0003], monthly maximum [95%CI -10.0125 to -2.07884; P 0.0034] and minimum temperature [95%CI -6.77 to -1.838; P 0.0008] were found to have a significant relationship with distribution of EVD emergence. From 2001 to 2018, the area of EVD emergence lost 2.25Mha of tree cover, equivalent to a 7.2% decrease in tree cover since 2000. MVD distribution was also observed to have an inverse relationship with minimum temperature [95%CI -6.7728 to -1.8380; P 0.0008] and a direct association with maximum temperature [95%CI 1.8275 to 8.0317; P 0.0018] and total precipitation [95%CI 0.015921 to 0.06010; P 0.001]. From 2001 to 2018, the regions contributing to the emergence of MVD lost 593kha of tree cover, equivalent to a 9.6% decrease in tree cover since 2000.

**Figure 1:**
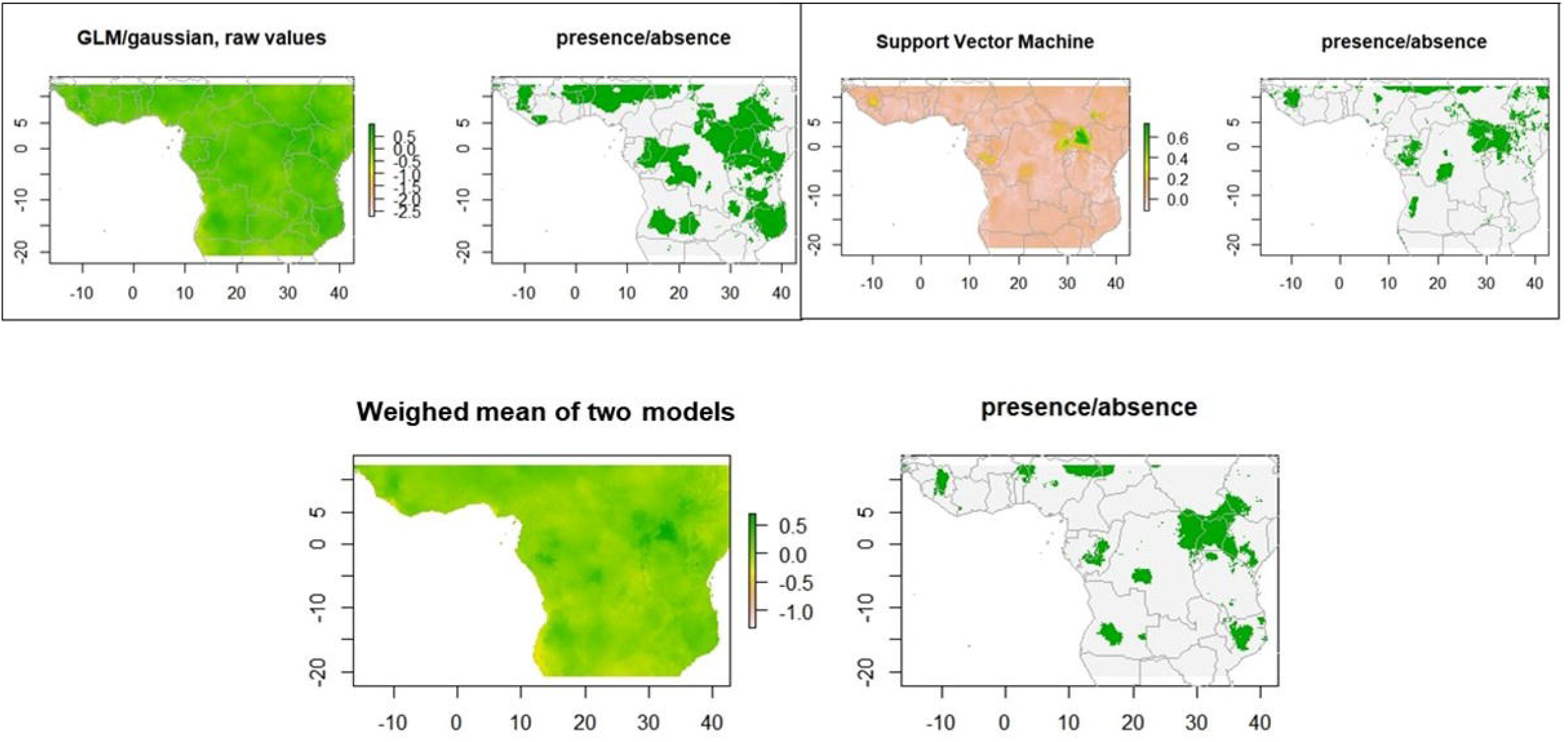
Map illustrating the predictive risk of Ebola viral disease using generalized linear models (GLM), support vector machine (SVM) and the weighed composite models.

**Figure 2:**
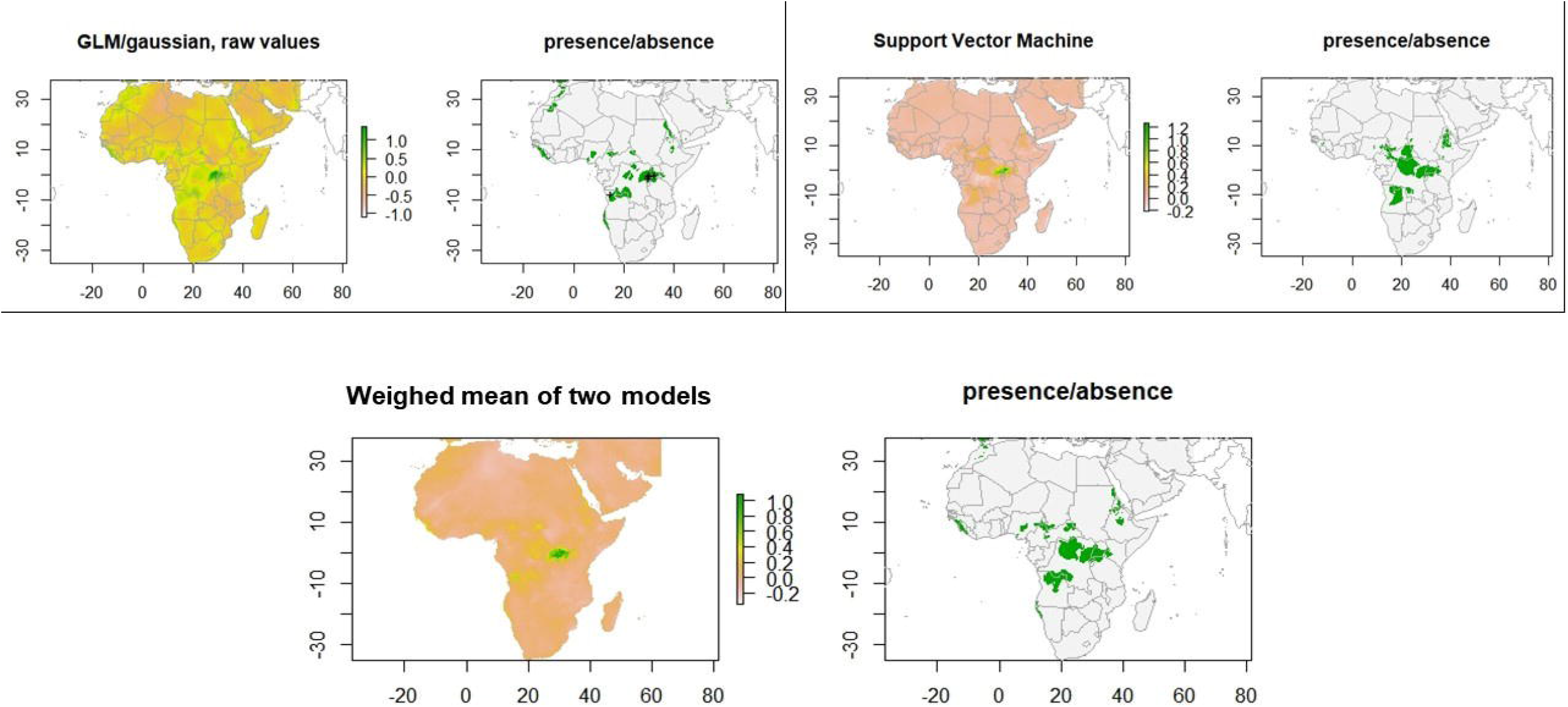
Map demonstrating the predictive risk of Marburg viral disease using generalized linear models (GLM), support vector machine (SVM) and the weighed composite models.

### Nipah virus disease

Elevation [95%CI -6.0114 to -1.4460; P 0.0014] along with the average maximum temperature [95%CI -8.5197 to -3.9108; P <0.0001] were found to have a negative correlation while the average minimum temperature [95%CI 2.6843 to 8.5860; P 0.0002] had a positive association with the distribution of Nipah disease (Figure 3). The occurrence sites in Bangladesh are comprised of the Lower Gangetic Plains Sundarbans mangroves vegetation and no intact forest.

**Figure 3:**
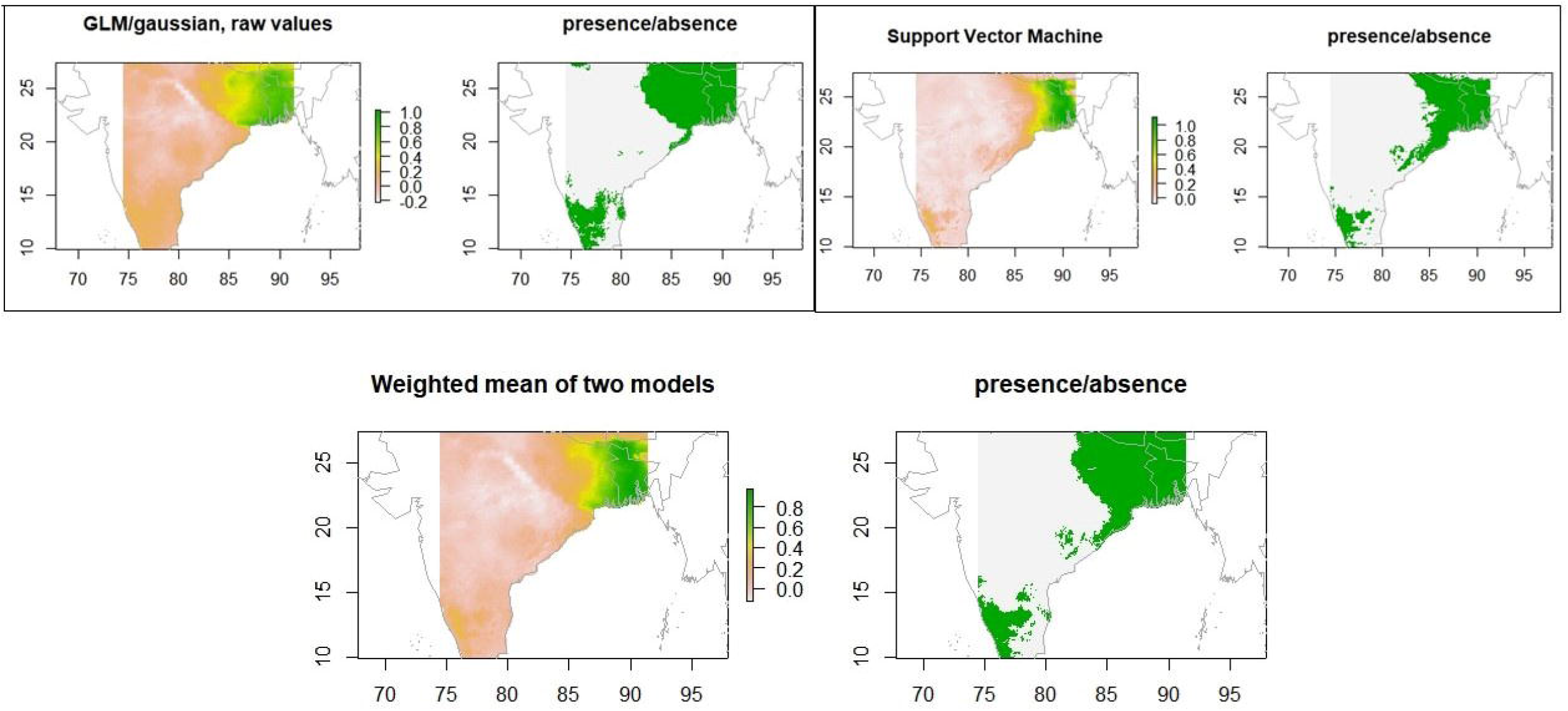
Map illustrating the predictive risk of Nipah using generalized linear models (GLM), support vector machine (SVM) and the weighed composite models.

However, between the years 2016 and 2018, Kerala (Figure 3) lost 23.2kha of tree cover, equivalent to a 0.89% decrease in tree cover since 2000.

### Rift Valley Fever

We found no significant relationship between the distribution of the RVF emergence (Figure 4) and the regions of cattle and goat grazing. However, we observed significant correlations with the average minimum temperature [95%CI 0.05881 to 0.14845; P <0.0001]. The tree cover loss in the regions with RVF occurrence is negligible.

**Figure 4:**
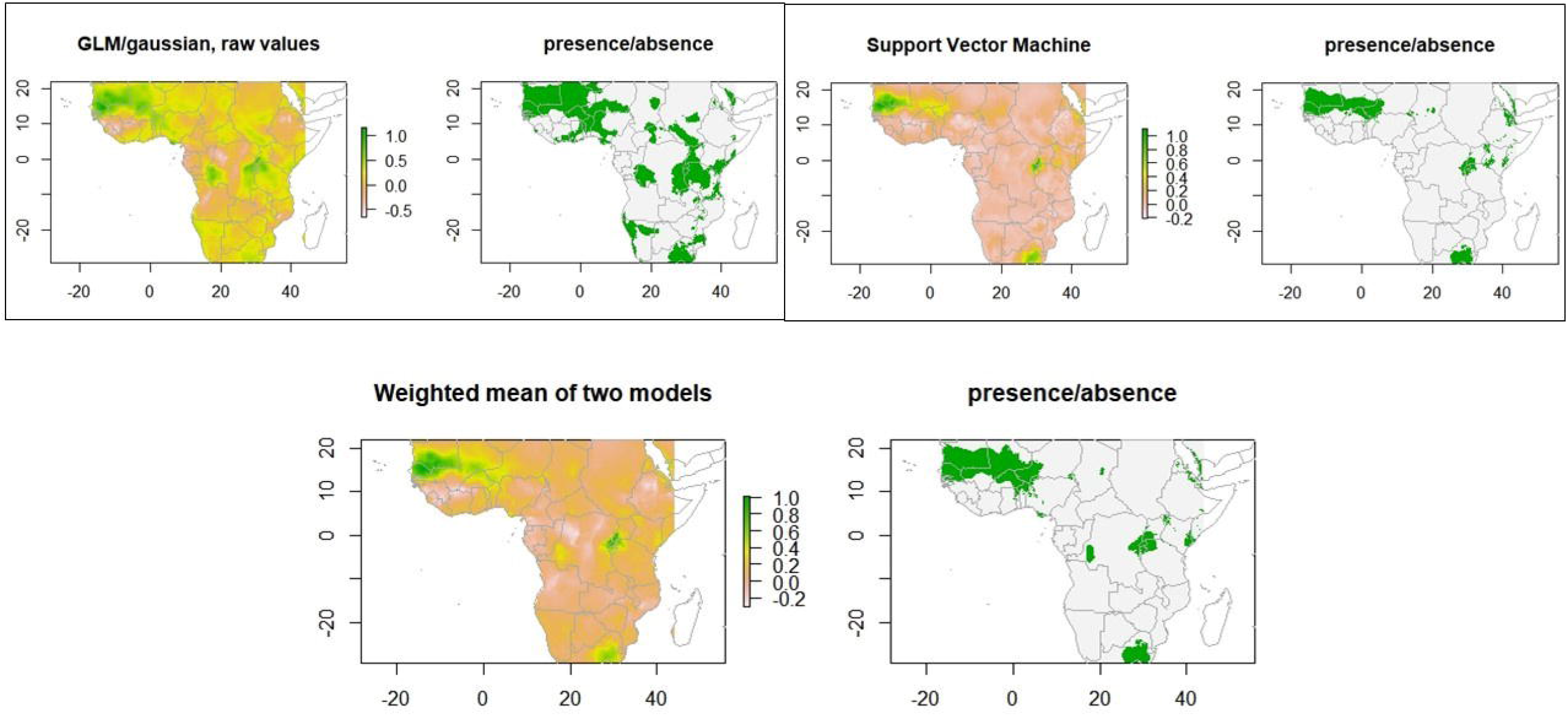
Map demonstrating the predictive risk of Rift Valley Fever using generalized linear models (GLM), support vector machine (SVM) and the weighed composite models.

### Coronavirus diseases

Elevation [95%CI -0.0014 to -0.0007; P <0.0001] and the average minimum temperature [95%CI 0.0067 to 0.0218; P 0.0003] had a positive association with the distribution of coronavirus diseases (Figure 5). From 2001 to 2003, Guangdong, the province where SARS emerged, lost 83.3kha of tree cover, equivalent to a 0.96% decrease in tree cover since 2000. The regions involved MERS had a negligible loss in tree cover.

**Figure 5:**
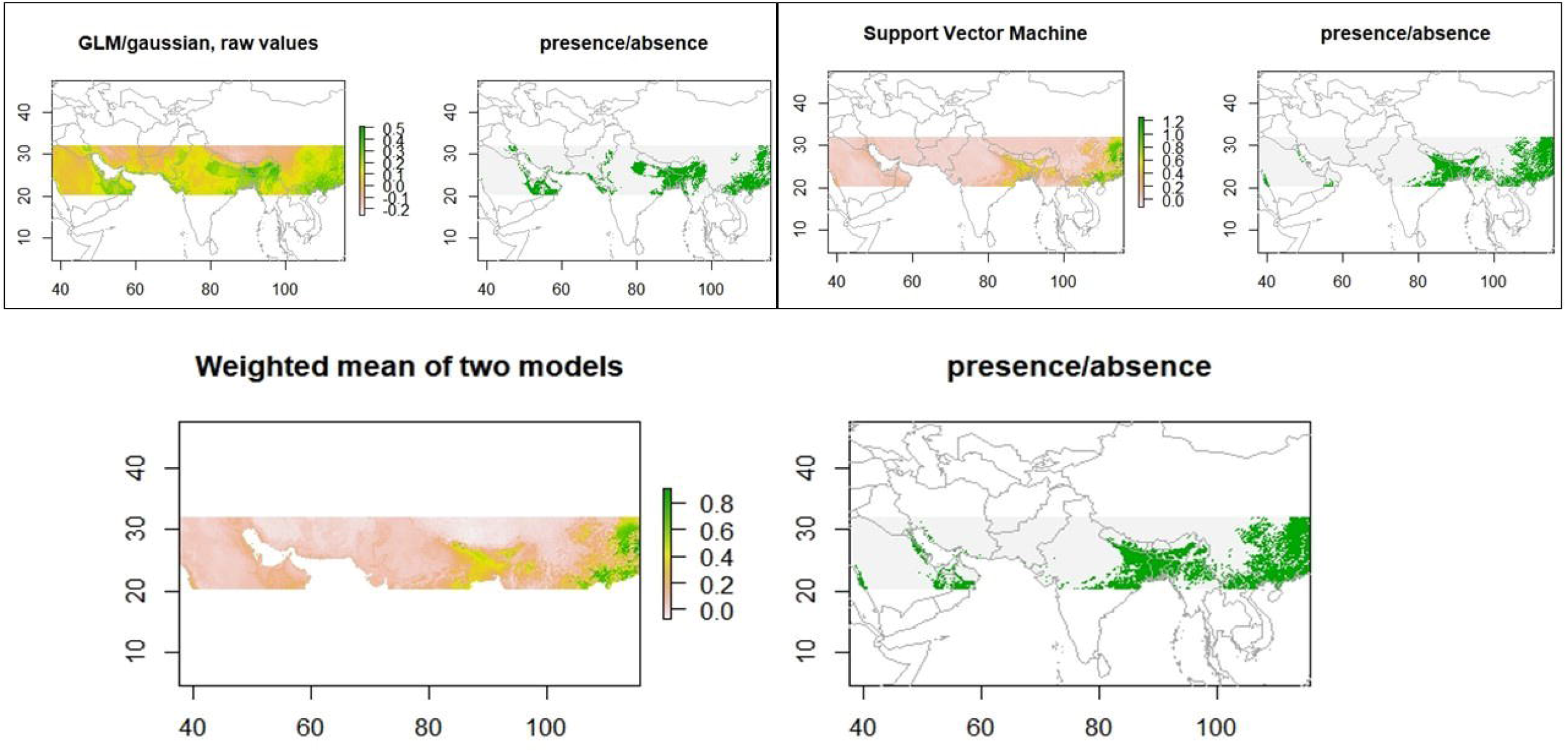
Map illustrating the predictive risk of Coronavirus diseases – SARS and MERS using generalized linear models (GLM), support vector machine (SVM) and the weighed composite models.

### Disease X

The weighed means of the AUC from the BPD models were used to create a threshold and the potential hotspots for the emergence of disease X was mapped (Figure 6).

**Figure 6:**
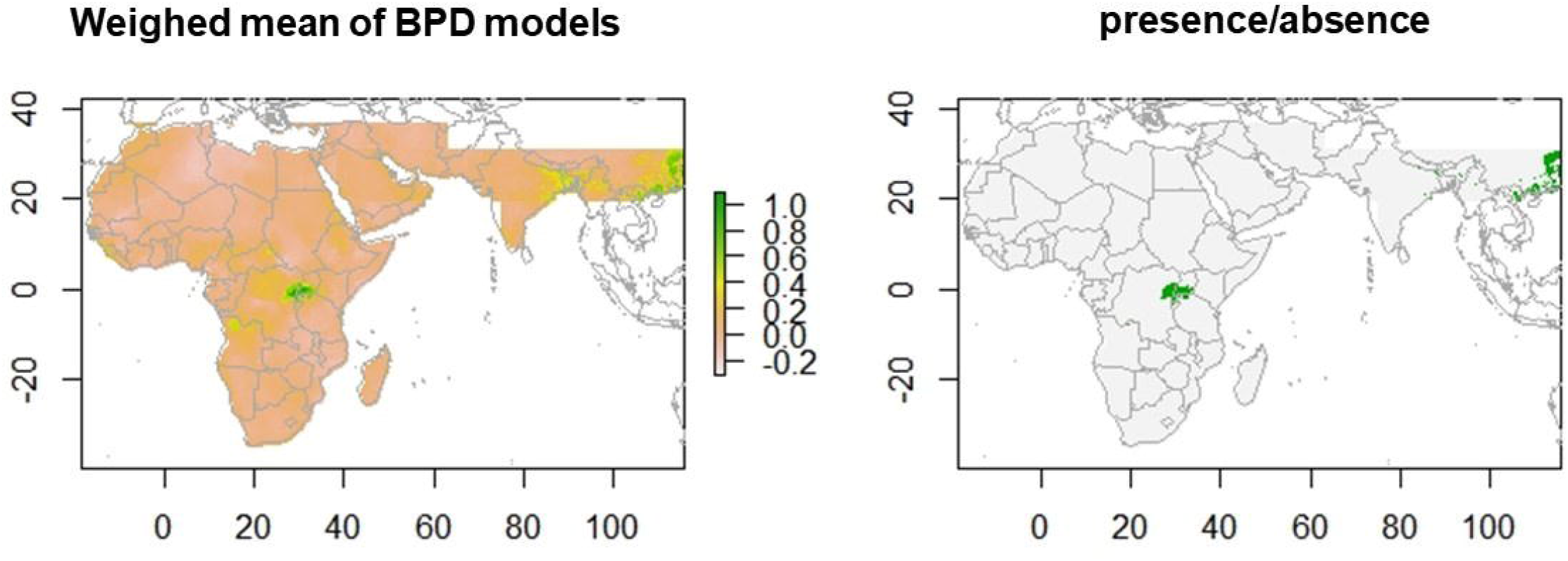
Map illustrating the potential hotspots of Disease X based on the weighed composite models of the blue print diseases list (BPDs).

## Discussion

Our study was the first to map the predictive risk of the disease emergence using species distribution models and to establish the impact of environmental factors on their emergence. We also identified the potential hotspots for future emerging infectious diseases based on the predictive models of BPDs. The results of our study have shown that the disease emergence is spatially dependent on the bioclimatic factors such as elevation, tree cover loss and climatic covariates. These factors could thus be utilized to identify and predict the hotspots of Disease X.

We found elevation to have a significant influence on the distribution of Nipah and coronavirus diseases. Studies have hypothesized that Nipah emergence in lower Gangetic plains and at the low-lying backwaters could be attributed to flooding, which causes destruction of mammal habitats(17). Rapid changes to habitats lead to starvation and migration of the known reservoirs of the Nipah virus, the bats, with contamination of fruit trees near human dwelling and increased exposure to the pathogen. Our results confirm this hypothesis associated with low-lying plains, flooding and emergence of Nipah disease. Coronaviruses are also linked with bat reservoirs. We hypothesize that similar events leading to destruction of bat habitats due to flooding at the low-lying regions such the wadis near Jeddah and Zhujiang Basin at the Province of Guangdong triggered the emergence of MERS and SARS respectively.

A recent study found that the increase in surface temperature and the unpredictability in the seasonal rainfall due to climate change, has an indirect effect on disease emergence through the sudden changes to the reservoirs habitats, biodiversity loss and small mammal migration (*18*). Our results demonstrate that increase in the residual temperature also known as the minimum temperature has a direct influence on the emergence and distribution of the BPDs. This direct spatial dependence of disease emergence on minimum temperatures is worrying. With climate change, the increasing night minimum temperatures lengthens the freeze-free season in most mid- and high latitude regions (*19*). These conditions with worsening climate change could increase the potential latitudinal extent of disease emergence. This hypothesis has been established from the vector-borne BPDs emergence predictions like CCHF and Zika (*5,20*).

Our study observed that deforestation is another important factor driving the emergence of BPDs. Studies have established the impact of deforestation and migration of bats on the occurrence of EVD, Nipah and SARS diseases (*11, 21, 22*). Most of the models showed tree cover loss at 100km around the occurrence of the BPDs (see for example EVD, Figure 7).

**Figure 7:**
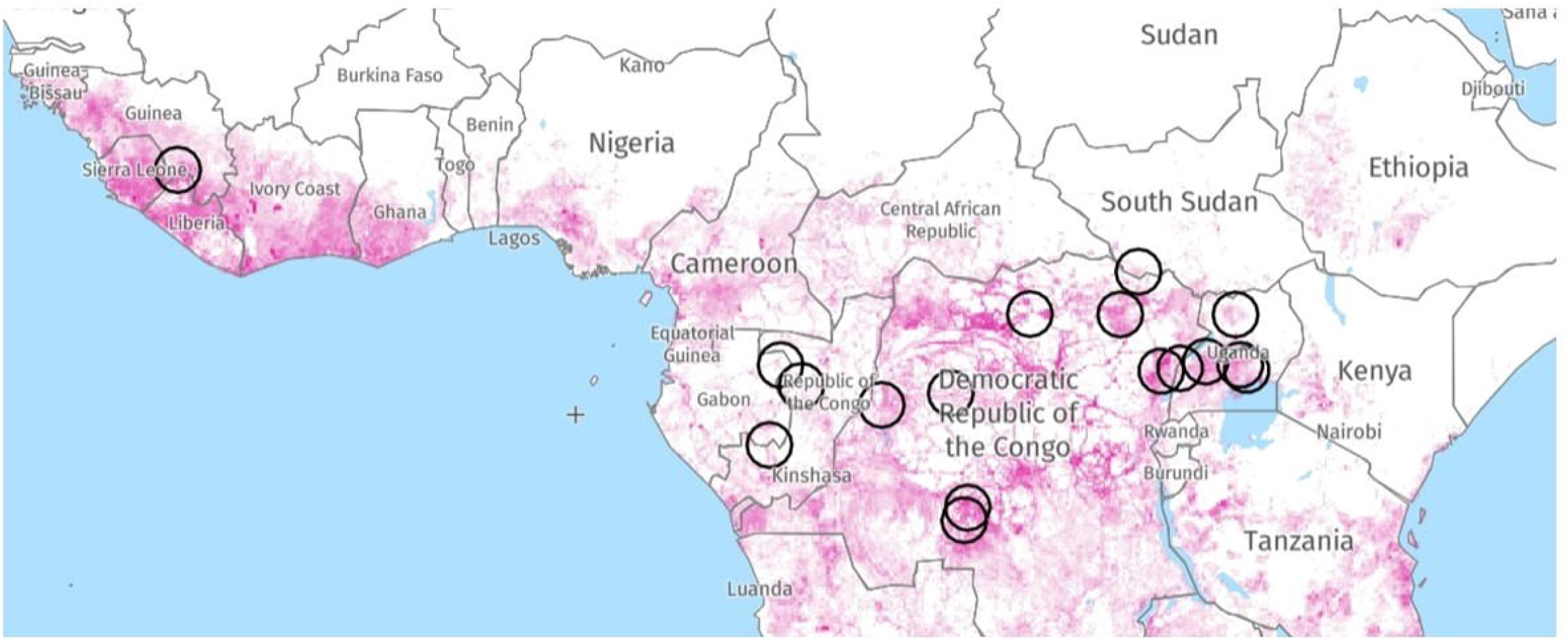
Tree cover loss around the occurrences of Ebola virus disease (EVD), spatial buffer of 100km, between the years 2001-2018.

Biodiversity loss, due to anthropogenic changes to their habitat such as deforestation, land fragmentation for agriculture, increase in demand for protein and climate We hypothesize that a decrease in predators could cause an imbalance in the natural predator-prey equilibrium leading to an increase in small mammal reservoirs and thus to viruses transmission via disease vectors such as ticks. Lesser species diversity and inter-species interactions makes spillover easier towards the accidental human hosts. This has been termed as the “dilution effect” (*23*).

As with all mathematical models, our study has its limitations. The size of the environmental predictor raster layers limited our spatial extent of the models. We chose the quality of the satellite imagery and resolution over a global perspective with poor or outdated data. Our study does not have a temporal component in the form of times series, which would be interesting especially with the climatic covariates. We mitigated this by choosing recent raster data corresponding to the period of the study and linking the spatiotemporal presence and pseudo-absence points to corresponding climatic monthly covariates. Despite these limitations, our study is the first to confirm the validity and effectiveness of using SDMs and other mathematical models to predict and identify the potential hotspots for BPDs. The use of a biogeographic approach in disease modelling offers a wider perspective on the environmental drivers and highlights the importance of climate change in the context of disease emergence. Most of all, our study results observed that the potential hotspots for an unknown disease X is located in Uganda and China (Figure 6). It is interesting to note that the associated predictive risk map includes the region of Wuhan, the epicenter of the ongoing COVID-19 outbreak.

## Conclusion and recommendations

In our study with the use of a biogeographic approach, satellite imagery and SDMs, we were able to identify Wuhan as a potential hotspot of disease emergence in the absence of COVID-19 data. We confirm that distribution of disease emergence in humans is spatially dependent on environmental factors such as landscape, tree cover loss and climatic covariates. The following are our recommendations to get ahead of the curve in the prediction and prevention of disease emergence:

1. Evaluate regions at high risk of flooding and identify them as hotspots for disease emergence at the tropics. With the unpredictability in rainfall and rising sea levels, these regions are in needs of active disease surveillance.
2. The direct relationship of the disease emergence with rapid changes in surface temperatures pose the threat of spatial latitudinal extension of the disease. Urgent need of global efforts to communicate the impact of climate change on future emergence of a disease like COVID-19 and thus to include EIDs when evaluation the economic costs of climate change.
3. Alternate solutions to deforestation, land fragmentation for livestock farming and bushmeat consumption to meet the growing protein demand.
4. The ongoing COVID-19 is not unprecedented but rather predictable using simple mathematical modelling techniques and freely available satellite imagery. We recommend using a biogeographic approach to predictive risk mapping to identify potential hotspots of disease emergence.

## Data Availability

The manuscript uses public data and the data sources are citated

